# Unhealthful plant-based diet associates with frailty risk predominantly in men with low income from the UK Biobank cohort

**DOI:** 10.1101/2024.07.23.24310856

**Authors:** Kerstin Schorr, Mar Rodriguez-Girondo, Niels van den Berg, Lisette CPMG de Groot, P. Eline Slagboom, Marian Beekman

**Affiliations:** Department of Biomedical Data Sciences, Leiden University Medical Center, Einthovenweg 20, 2333 ZC Leiden, The Netherlands; Division of Human Nutrition and Health, Wageningen University, Wageningen, P.O. Box 176700 AA Wageningen, The Netherlands

**Keywords:** plant-based diet, frailty, socio-economic status, health inequality, sex differences, dietary pattern

## Abstract

**Objective:** Plant-based diets (PBD) are generally promoted as beneficial for health. However, whether this is also the case at older ages, when energy deficits, muscle loss and frailty affect health, is unclear. Research has shown that among older adults, particularly in men, a healthful PBD is associated with a lower frailty risk. This relation was however, not studied in the context of socio-economic status (SES), a major factor influencing the risk of frailty. Therefore, we aim to assess whether plant-based diets associate with frailty risk at older ages and whether this association is moderated by sex and income in a large population-based dataset.

**Methods:** we investigated data from the UK Biobank (UKB) (n=73 180, mean age=55.48±7.87). We applied a plant-based diet index [range 17-85], differentiating between a healthful (hPDI) and unhealthful plant-based diet (uPDI). Frailty was assessed by the Fried frailty phenotype and categorized into 0-4 symptoms of frailty. Average household income was divided into three categories: low (<18.000 £), medium (18.000-52.000 £) and high (>52.000 £). We applied an ordinal logistic regression model with frailty as the categorical outcome and PDI as continuous predictor while adjusting for age, sex, ethnicity, education, BMI and UKB assessment center. Secondly, we included an interaction term (PDI*sex*income). To identify subgroups driving any interactions, we stratified by sex and subsequently by income group to determine the effect of PDI in subgroups while additionally adjusting for lifestyle factors.

**Results:** a 10-unit increase in hPDI, was associated with 3.4% lower odds for frailty (OR=0.966, 95%CI [0.946, 0.987]), whereas a 10-unit increase in uPDI was associated with 7.7% greater odds for frailty (OR=1.077, 95%CI [1.054, 1.101]). The association between uPDI and frailty was moderated by income and sex (uPDI*income*sex, p=0.002), whereas no such moderation was found for hPDI (p=0.602). Subsequent stratification reveals a significant effect of uPDI on frailty particularly among men with low income (OR=1.177, 95% CI [1.069, 1.298]), but not for women. This association in men largely persisted after adjustment for additional lifestyle factors (OR=1.119, 95%CI [0.995, 1.258]).

**Conclusion:** we observed that adherence to an unhealthful plant-based diet was associated with a higher risk for frailty. This relation was especially observed for men with lower incomes and not explained by other lifestyle factors. Therefore, this group may profit from refraining from an unhealthful plant-based diet.

**Highlights:**

- Healthful plant-based diet associated with lower frailty risk
- Association of unhealthful plant-based diet and higher frailty risk is moderated by income and sex
- Unhealthful plant-based diet is associated with frailty particularly in men with low income

## 1. Introduction

Plant-based diets, characterized by a high intake in plant foods and comparatively low intake of animal foods, have gained attention in recent years as a shift towards more plant foods is recommended by the EAT-Lancet committee [1] as well as dietary guidelines worldwide. Further, plant-based diets are recommended for the prevention of age-related diseases, in particular metabolic diseases. However, for vulnerable populations strict plant-based diets may also lead to energy deficits and inadequate protein intake, contributing to a loss of muscle mass, which poses a risk factor for frailty [2]. Frailty has been defined as increased vulnerability resulting from an age-associated decline in function and may put older adults at an increased risk for adverse outcomes, such as hospitalization and mortality [3]. Meanwhile, studies have shown that a healthful plant-based diet may actually lower the risk for frailty, although not in both sexes equally [4, 5]. The role of diet in the prevention of frailty, may be explained by anti-inflammatory properties of plant foods [4, 6] and men in particular may respond more to the effects of an inflammatory diet [6, 7]. At the same time, previous research has shown that women are generally more likely to become frail over the life course [8], indicating differences between men and women exist in the prevalence of frailty and response to lifestyle factors [9]. It is therefore important to further investigate the interplay of diet and sex on frailty risk.

Sex-differences also exist regarding socioeconomic background and its effect on frailty. Higher socioeconomic status (SES) has been linked to a healthier lifestyle and reduced frailty risk, while low SES groups have benefitted more from improving their lifestyle [10]. In addition, sex differences were observed: women’s frailty risk was determined more by education, whereas in men income had a stronger influence [11]. Similarly, individuals with a lower SES and an unhealthy lifestyle have been found to have a higher mortality risk compared to individuals with high SES and a comparable unhealthy lifestyle, with men with low income being a particular at-risk group [12]. Thus, these findings point towards a potential interaction of lifestyle, sex, and income. It is currently not known whether each of these characteristics associate additively and independently with the development of frailty, or whether the interplay of these factors impacts frailty risk.

We therefore aim to identify at-risk groups for frailty which may be target groups for future dietary and lifestyle interventions. We focus on the question whether plant-based diets are differently associated with frailty in men and women and whether this association dependent on income. To quantify adherence to a plant-based diet, we apply a plant-based diet index, differentiating between adherence to a healthful and unhealthful plant-based diet to a population of 73,180 adults (mean age=55.48 years) from the UK Biobank.

## 2. Methods

### 2.1. Study population

For this study, we used data from the UK Biobank, a multicenter study with >500,000 participants conducted in England, Scotland and Wales [13]. Baseline measurements were carried out between 2006 and 2010. For the purpose of this study, we included participants that had baseline data on all variables of interest available and filled at least two 24h-diet recalls. Our final sample consisted of n=73,180 (mean age=55.48 years) (Table 1).

**Table 1:**
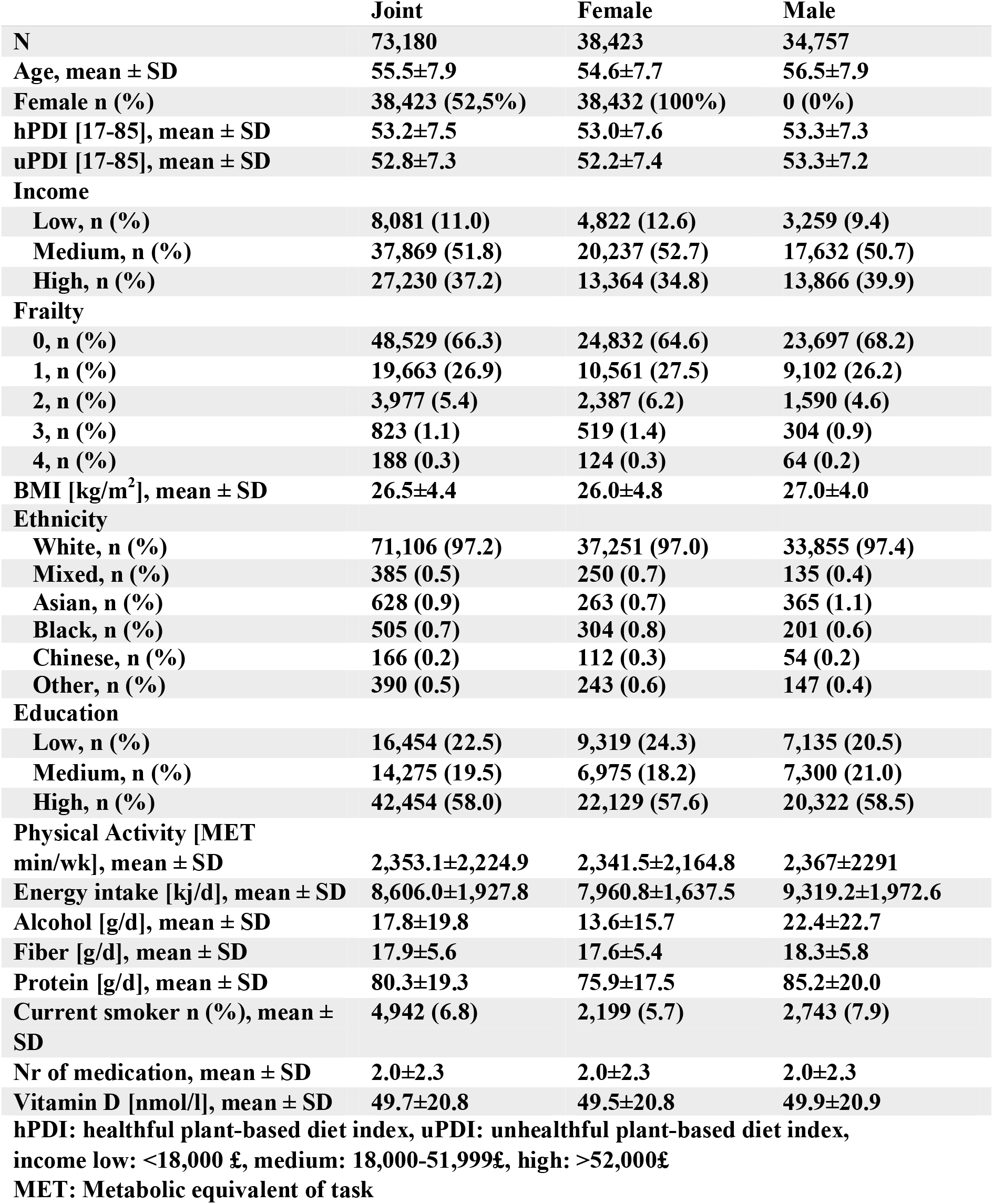
Descriptive statistics over UK Biobank study population.

### 2.2. Plant-based diet index

For the calculation of the plant-based diet index, we used preprocessed data that provided information on dietary intakes of 93 food groups in grams per day (Category 10018, Suppl. Table 1). This dataset was created by Piernas et al. in 2022 based on raw data from the Oxford WebQ tool [14, 15]. In this questionnaire, participants indicated their intake in serving sizes within the last 24h. In our study, participants that completed at least two questionnaires were included in the analysis, to gain an estimate of usual intake. We further excluded participants with unrealistic energy intake as described previously [16]. To calculate the PDI for each participant from this data set, intakes of 17 food groups, consisting of healthy plant foods (e.g., wholegrains, vegetables, nuts), unhealthy plant foods (e.g., sweets, refined grains) and animal foods, were averaged across questionnaires (Suppl. Table 2). Vegetable oil was not included since insufficient information was available. The PDI was calculated following standard procedure [17]: The intakes per food groups were divided into sex-specific quintiles. For the healthful plant-based diet index (hPDI) this includes scoring healthy plant foods positively, whereas unhealthful plant foods were scored negatively, and animal foods were always scored negatively. Since intakes in some food groups were low, the cut-off for several quintiles was 0. Therefore, quintiles with a low intake (0 g until the cut-off of the first non-zero quintile) were summarized and awarded a 1. The remaining quintiles were then scored as normal, i.e., those above the fifth quintile receiving a score of 5 etc. The final score has a theoretical score ranging from 17-85, with higher scores indicating greater adherence to a healthful (hPDI) or unhealthful (uPDI) diet, respectively. The index was adjusted for energy intake using the residual method [18].

### 2.3. Frailty Index and Income

The frailty index is an adaption from Fried’s frailty index with variables found in the UK Biobank [19, 20]. This adapted Fried’s frailty index includes hand grip strength, usual walking pace, weight loss in the past year, feelings of exhaustion in the past week and physical activity in the last 4 weeks. According to this definition participants were considered frail if they fulfil at least three of the following criteria: weight loss in the previous year, tired or low energy on more than half the days in the past two weeks, none, or light physical activity with a frequency of once per week or less, slow usual walking pace and low grip strength (according to sex and bmi specific cut-offs). Participants that met one or two criteria were classified as pre-frail. The process has been described in more detail elsewhere [20]. Since only a small percentage of our sample showed all 5 symptoms of frailty (n= 21), those who show 4 or 5 symptoms of frailty were grouped together (n=188).

Total household income before tax was collected in 5 categories. For the sake of data analysis this variable was coded into three categories: low (<18,000£), medium (18,000 – 51,999£), and high (>52,000£).

### 2.4. Statistical analysis

We first assessed whether all variables of interest, i.e., PDIs, frailty and income were independently associated with each other. To assess the association between plant-based diet adherence and frailty, we applied an ordinal logistic regression model with frailty as the categorical outcome variable and PDI as continuous predictor (models 1a and 1b). For determining the association between frailty and income, an ordinal logistic regression model was fit with frailty as the outcome and income as the categorical predictor (model 2). Lastly, an ordinal logistic regression model was fit with income as the categorial outcome variable and PDI as a continuous predictor (models 3a and 3b). All models were adjusted for age, sex, bmi, ethnicity, and education, due to their relevance in the development of frailty [8, 21-23]. In addition, the analyses were adjusted for the assessment center in which data was collected to take into account potential differences between centers.

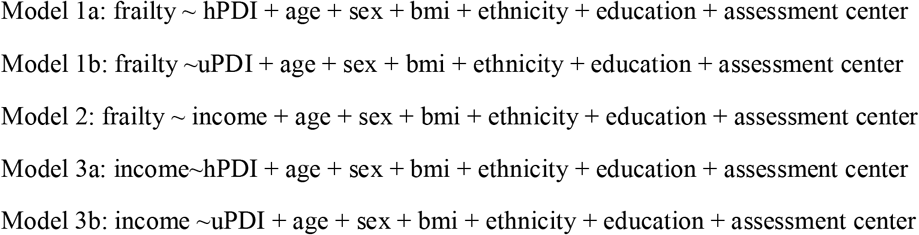

In a second step, we aimed to address the question whether there is an influence of sex and income on the association between frailty and hPDI (model 4a) or uPDI (model 4b), respectively. For this, we added a three-way interaction term of sex, hPDI/uPDI and income to an ordinal logistic regression model with frailty as the categorical outcome variable.

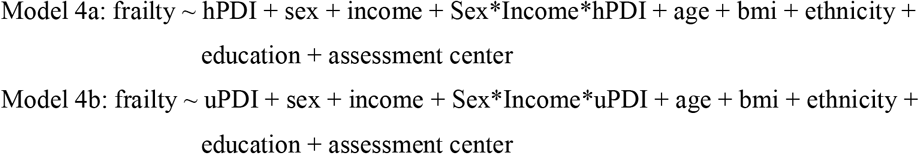

To assess difference in effect between men and women, we stratified the interaction models by sex.

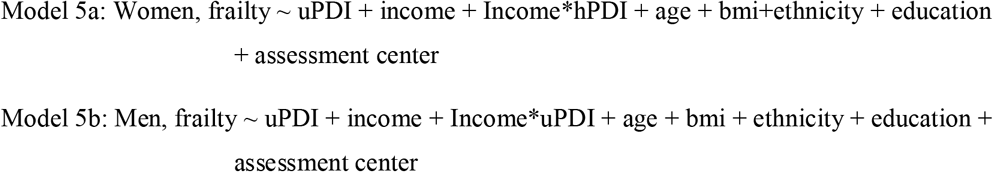

To identify potentially explanatory characteristics for the interaction, we further adjusted for lifestyle and health-related factors (physical activity, alcohol, fiber and protein intake, vitamin D levels, smoking status, number of prescribed medications). Low physical activity has been previously identified as a risk factor for frailty [24]. Alcohol can be a risk factor for a multitude of diseases, and multimorbidity is a risk factor for frailty [25]. For the same reason we adjust for number of prescribed medications as an indicator of overall health status [20]. Vitamin D is related to bone health and metabolism and may therefore also play a relevant role in the development of frailty [26]. Both, fiber, and protein intake have been associated with increased muscle mass and may therefore be protective for frailty [27, 28]. Alcohol, fiber and protein intake were adjusted for energy intake via the residual method [18].

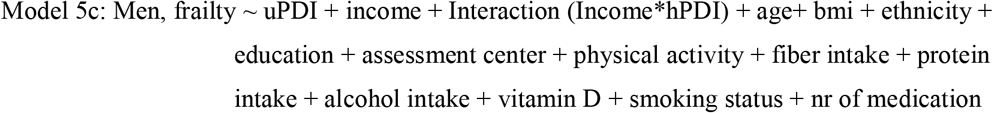

In the last step, we stratified models by income group for the models that showed a significant interaction effect of PDI and income in the previous step to identify the effect of uPDI on frailty risk per subgroup.

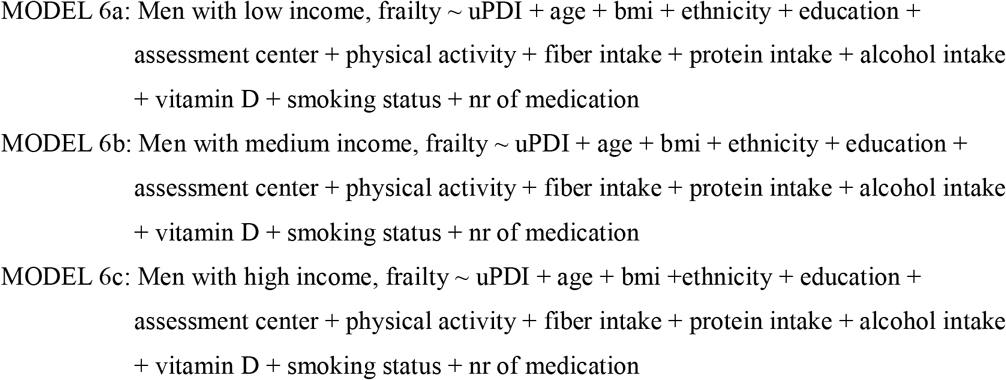

We carried out a sensitivity analysis in which we further adjusted for atypical dietary intake. When filling the 24h diet records, participants could indicate whether they followed a typical diet that day. We calculated the percentage of questionnaires with atypical intake and added this to our models. We further replicated all analysis with a binary logistic regression (i.e., binarization of frailty in no symptoms of frailty (scores 0), and mild to severe frailty (scores 1-5) to validate to our findings. All analysis were carried out in R 4.3.2.

## 3. Results

The complete sample consisted of n=73,180 mostly white adults with a mean age of 55.5 years. A slight majority of the sample consisted of women (52.50%). Women had slightly lower adherence to uPDI, were more likely to be in the low-income group compared to men. Further, women had a lower BMI, consumed less alcohol and were less likely to be smokers than men. Despite their seemingly healthier lifestyle choices, women reported higher medication intake and were more likely to be frail (Table 1).

### 3.1. Plant-based diet and income are independently associated with frailty risk

We first assessed whether frailty is (univariately) associated with PDI (model 1a and 1b) and income (model 2), and whether PDI is associated with income (model 3a and 3b). Both, hPDI and uPDI were associated with frailty, as expected in different directions: per 10-unit increase in dietary index, higher hPDI was associated with 3% (OR=0.966, p=0.002) lower odds for frailty, whereas uPDI was associated with 8% (OR=1.077, p<0.001) greater odds for frailty (Suppl. Table 3).

High income, while adjusted for covariates, such as education level, was associated with 44% (OR=0.558, p<0.001) lower odds for frailty, whereas those in the medium income group had 28% (OR=0.718, p<0.001) lower odds (Suppl. Table 4). Lastly, we observed a 10-unit increase in hPDI and uPDI to be associated with 7% (OR=0.928, p<0.001) and 3% (OR=0.969, p=0.003) lower odds for high income, respectively (Suppl. Table 5).

### 3.2. Unhealthful plant-based diet, income and sex have interactive effect on frailty risk

We observed differences in the distribution of PDI, income and frailty risk between sexes in our study population (Table 1). To assess whether the overall association between PDI and frailty is moderated by income, we included an interaction term between PDI, sex and income.

We observed that for hPDI, the three-way interaction terms, i.e., “hPDI-middle income-male sex” and “hPDI-high income-male sex” showed no significant results (model 4a+b, Suppl. Table 6). Interestingly, we did find a significant interaction between high and middle income, male sex and uPDI (uPDI-high income-male sex: coef=-0.023, p=0.002; uPDI-middle income-male sex: coef=-0.016, p=0.024, Suppl Table 6).

### 3.3. Effect of unhealthful plant-based diet primarily in men of low income

To further pinpoint the interaction effects of sex, uPDI and income on frailty, we performed sex-stratified analyses (model 5a+b). In women uPDI was not associated with frailty and neither did it interact with income (Suppl. Table 7). Meanwhile, in men, we observed an association of uPDI with frailty (OR=1.249, p<0.001) with a significant interaction between uPDI and medium (p=0.007) and high income (p<0.001).

The interaction observed in men may be explained by lifestyle and health-related factors such as physical activity, smoking, alcohol consumption and vitamin D levels. Therefore, we further adjusted the association for these additional lifestyle and health-related factors (model 5c). Interestingly, the interaction term between uPDI and income remained significant, indicating there is a difference in the association between uPDI and frailty by income which is not explained by lifestyle and health-related factors tested (model 5c, Suppl. Table 8). Subsequently, to visualize the interaction effect, we further stratified by income group (models 6a-c). We observed that for men with a high income, uPDI was not associated with frailty (OR=0.994, 95% CI [0.933, 1.059]), while in contrast every 10-unit increase in uPDI was associated with 18% (OR=1.177, 95% CI [1.069, 1.298]) greater odds for frailty in men with a low income (Fig. 1). This association persisted after adjustment for additional health- and lifestyle factors (OR=1.119, 95%CI [0.995, 1.258], Fig. 1).

**Figure 1.**
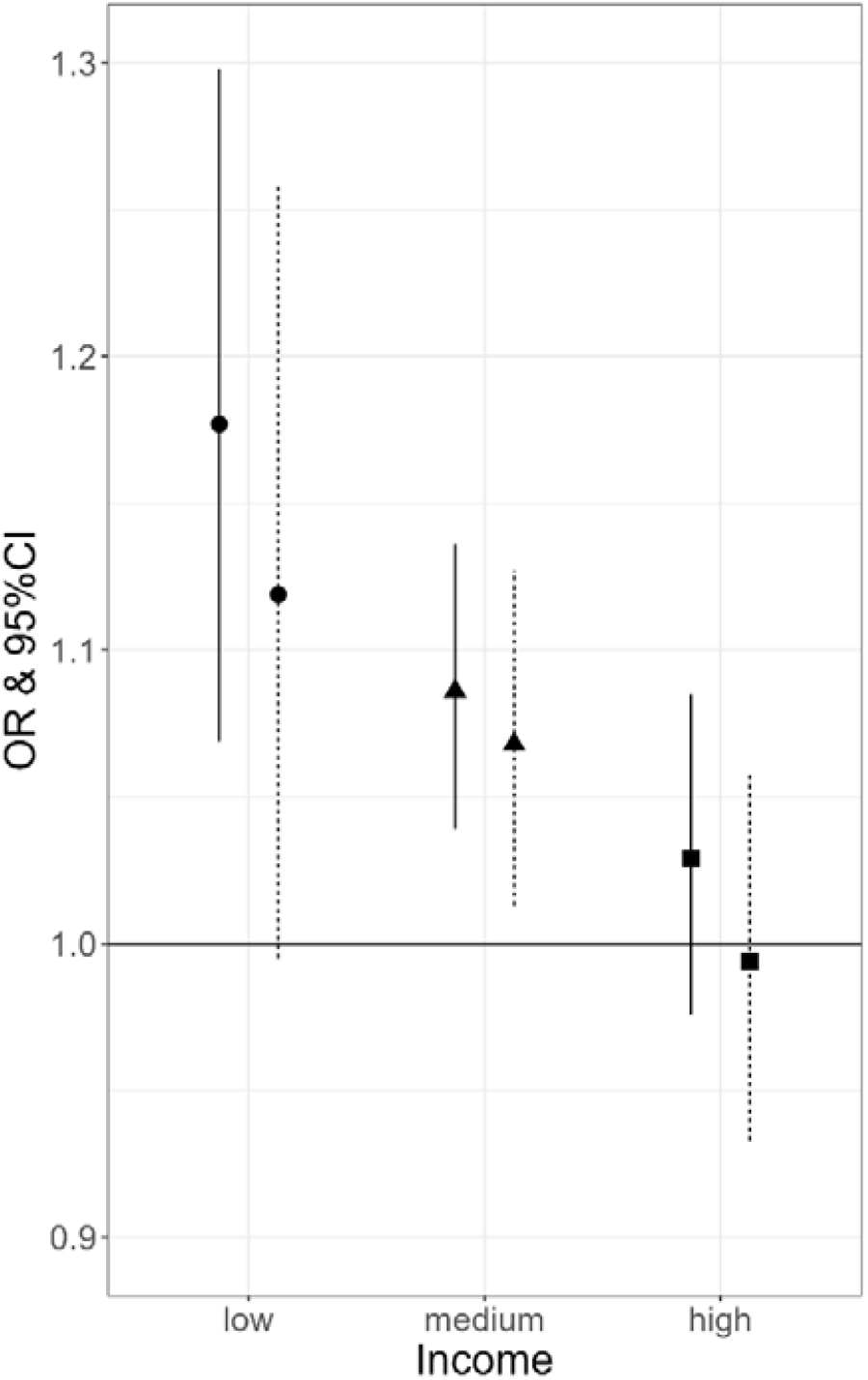
Association between unhealthful plant-based diet index (uPDI) and frailty in men of different income groups. OR per 10-unit increase in uPDI − Basic model --- fully adjusted model ● low income ▲ medium income ■ high income

### 3.4. Sensitivity Analysis

To test the influence of atypical dietary intake, we adjusted for the percentage of diet questionnaires in which an atypical diet was reported. We found that atypical intake was not associated with the overall results (Suppl. Fig 1). Further, replication of our results with a binary logistic regression yielded similar results (Suppl. Fig 2).

## 4. Discussion

In a large dataset of over 70 thousand individuals, we dissected how plant-based diets, sex, and income associate with frailty. We observed for both men and women that greater adherence to a healthful plant-based diet was associated with a lower risk of frailty, and greater adherence to an unhealthful plant-based diet was associated with higher risk of frailty. In addition, we established that higher income and male sex were associated with lower risk of frailty. Because sex, income and diet are known related factors [4, 8, 11], we tested for a three-way interaction of these variables in their association with frailty. While we observed no interaction for hPDI, sex and income on their effect with frailty, we did observe interaction of uPDI, sex and income on frailty. Specifically, men with low income and a high adherence to unhealthful plant-based diet had remarkable higher risk for frailty, which was not explained by physical activity, alcohol consumption or smoking. Remarkably, no effect of uPDI on frailty was observed in high income men or women in either income group.

Our findings, that a healthful plant-based diet was associated with a lower frailty risk, while an unhealthful plant-based diet was associated with a higher risk for frailty, is supported by other research [4, 5, 7]. Our observations of an interaction between income and unhealthful dietary pattern were also in line with previous research. A study in the UK Biobank concluded that the association between unhealthy lifestyle behaviors (including e.g. diet, physical activity, sleep duration and smoking) and adverse health outcomes were stronger among deprived populations [29]. Similarly, a more recent large population-based study reported that a healthy lifestyle benefited low socio-economic status (SES) populations more by having a stronger association with mortality risk compared to groups with high socio-economic status [12] In Chinese older adults a protective effect of high income on frailty was reported [11]. Our study is unique in that we investigate the interplay of all three factors, income, sex, and plant-based diet adherence combined, on frailty risk. We identified men with low income as an at-risk group for frailty when consuming an unhealthful plant-based diet, even adjusting for lifestyle factors, suggesting a unique effect of diet on frailty.

Several hypotheses have been proposed for the higher morbidity observed especially in men with low income. First, men are generally assumed to engage more in unhealthy behaviors [30]. Similarly, low SES groups are more likely to engage in unhealthy behaviors in extreme ways compared to higher SES groups. This has been discussed for alcohol consumption, with low income groups being more likely to engage in excessive drinking, but may also extend to other health behavior such as diet [31]. When comparing uPDI scores of the frailest individuals, we observe that, men with a low income had a higher uPDI (55.05) than high income men in the same frailty category, suggesting men with low income engaged more in unhealthy dietary habits (50.43). While a clear explanation for our findings is lacking, we observed that women in our sample had consistently higher intakes of vegetables, fruit, tea, and coffee as well as dairy compared to men. All these food groups have been associated with a lower frailty risk, supposedly due to concomitant higher intakes of antioxidants, protein, and calcium [32-34]. Further, men with low income and frailty were the subgroup with the highest intake of soda (data not shown), potentially explaining the association with frailty in this population group [35]. Future research may explore how dairy and a reduction of soda consumption may fit into a healthful plant-based diet to contribute to lower frailty risk.

At the same time research suggests that low income groups have less access to the medical system or are less inclined to make use of medical care [29]. Further, low income often goes hand in hand with increased stress and mental burden associated with the challenge to make a living. In men, the mental distress of low socioeconomic status may intersect and be amplified by societal expectations to provide, leading to a greater importance of income in this group [36]. Men with a low income may therefore be a particularly vulnerable group and may be targeted by dietary interventions.

Surprisingly, we observed no interaction effect of income and plant-based diet adherence with frailty risk in women. Women are generally more likely to be frail yet have a lower mortality risk. This is known as the male-female health-survival paradox [8]. It has been proposed, that in women, other factors contribute more to frailty, shifting the focus away from lifestyle. For example, women accumulate more disabling conditions, such as arthritis, which can limit physical functioning [6, 37]. It has also been suggested, that inflammation may contribute more to the development of frailty in women [38]. At the same time, men have been proposed to benefit more from an anti-inflammatory diet, potentially contributing to lower frailty risk [39]. Previously, differences in the gut microbiome between sexes has been suggested as a potential mechanism [6], however more studies are needed to investigate the interplay of sex and frailty, the gut microbiome and risk factors such as diet and inflammation.

Our analysis has several limitations. Firstly, the UK Biobank sample was still relatively young with a mean age of 55 (range 40-70 years) years. Subsequently, cases of frailty, especially those who showed more than three symptoms of frailty, were relatively low in our sample and would be considered early frailty. Secondly, participants of the UK Biobank study tend to live in less deprived areas [40]. This is also supported by the fact that the majority of our sample had a medium or high income. In our analysis, we included participants who had filled at least two 24h diet recalls. It can be argued that two questionnaires may be not enough to accurately reflect usual intake. However, according to the median of questionnaires filled, we had 3 or more questionnaires available from 50% of our sample, a number that has previously been identified as sufficient for estimating energy intake [34]. While we adjusted for ethnicity, the vast majority of the sample was white, leading to limited transferability of our results to other ethnic groups. We did not assess the impact of biological factors, such as levels of inflammatory markers, the gut microbiome, or pre-existing conditions, although we approximated pre-morbidities by adjusting for medication intake. Further research may therefore extend our findings by taking into account these factors. Lastly, as we conducted a cross-sectional analysis, causality cannot be inferred and there is a possibility that reverse confounding contributed to the results. Our findings on the association between diet and frailty are however in line with other, longitudinal research, supporting our results. Future studies may replicate our findings in older, more deprived, and diverse populations and take into account markers of metabolic health.

The strength of our analysis lies predominantly in the large sample size of the UK Biobank making robust observations and allowing to explore the relative contribution of clustering factors influencing frailty. Despite potentially limited representativeness, it has been suggested, that exposure-disease relationships are still extrapolatable to the general population [40]. Further, the extensiveness of the UK Biobank database allowed us to adjust our analysis for a variety of lifestyle and health related factors. Additionally, the Oxford WebQ that was used to collect dietary data has been validated and shown to produce reliable results [14].

## 5. Conclusion

We conclude that adherence to an unhealthful plant-based diet contributes to frailty risk predominantly among men with low income (<£18k). We therefore identified an at-risk group, which may benefit from dietary and lifestyle interventions. Future research may focus on identifying potential dietary drivers of the identified association in older and more deprived populations.

## Data Availability

All data produced in the present study are available upon reasonable request to the authors

## Abbreviations

PBD: Plant-based diet
UKB: UK Biobank
PDI: Plant-based diet index
hPDI: healthful plant-based diet index
uPDI: unhealthful plant-based diet index
SES: socio-economic status

## 6. Acknowledgements

This research has been conducted using the UK Biobank Resource under Application Number 78275

## 8. Author Contributions

KS, MB, PES were involved in study concept and design, acquisition, analysis, and interpretation of data. NvB and MRG provided statistical and methodological advice. LCPGMdG provided critical input. KS performed statistical analysis and prepared the manuscript. All authors read and approved the final version.

## References

1. Willett, W., et al., Food in the Anthropocene: the EAT-Lancet Commission on healthy diets from sustainable food systems. Lancet, 2019. 393(10170): p. 447–492.

2. Cruz-Jentoft, A.J., et al., Nutritional strategies for maintaining muscle mass and strength from middle age to later life: A narrative review. Maturitas, 2020. 132: p. 57–64.

3. Clegg, A., et al., Frailty in elderly people. The Lancet, 2013. 381(9868): p. 752–762.

4. Maroto-Rodriguez, J., et al., Plant-based diets and risk of frailty in community-dwelling older adults: the Seniors-ENRICA-1 cohort. Geroscience, 2023. 45(1): p. 221–232.

5. Sotos-Prieto, M., et al., Association between the quality of plant-based diets and risk of frailty. Journal of Cachexia, Sarcopenia and Muscle, 2022. 13(6): p. 2854–2862.

6. Reid, N., et al., Sex-specific interventions to prevent and manage frailty. Maturitas, 2022. 164: p. 23–30.

7. Qi, R., et al., Plant-Based Diet Indices and Their Association with Frailty in Older Adults: A CLHLS-Based Cohort Study. Nutrients, 2023. 15(24).

8. Gordon, E.H., et al., Sex differences in frailty: A systematic review and meta-analysis. Experimental Gerontology, 2017. 89: p. 30–40.

9. Dorhout, B.G., et al., In-Depth Analyses of the Effects of a Diet and Resistance Exercise Intervention in Older Adults: Who Benefits Most From ProMuscle in Practice? J Gerontol A Biol Sci Med Sci, 2021. 76(12): p. 2204–2212.

10. Kheifets, M., et al., Association of socioeconomic status measures with physical activity and subsequent frailty in older adults. BMC Geriatrics, 2022. 22(1): p. 439.

11. Wang, H.-y., M. Zhang, and X. Sun, Sex-Specific Association Between Socioeconomic Status, Lifestyle, and the Risk of Frailty Among the Elderly in China. Frontiers in Medicine, 2021. 8.

12. Zhang, Y.-B., et al., Associations of healthy lifestyle and socioeconomic status with mortality and incident cardiovascular disease: two prospective cohort studies. BMJ, 2021. 373: p. n604.

13. Sudlow, C., et al., UK Biobank: An Open Access Resource for Identifying the Causes of a Wide Range of Complex Diseases of Middle and Old Age. PLOS Medicine, 2015. 12(3): p. e1001779.

14. Greenwood, D.C., et al., Validation of the Oxford WebQ Online 24-Hour Dietary Questionnaire Using Biomarkers. Am J Epidemiol, 2019. 188(10): p. 1858–1867.

15. Piernas, C., et al., Describing a new food group classification system for UK biobank: analysis of food groups and sources of macro- and micronutrients in 208,200 participants. European Journal of Nutrition, 2021. 60(5): p. 2879–2890.

16. Thompson, A.S., et al., Association of Healthful Plant-based Diet Adherence With Risk of Mortality and Major Chronic Diseases Among Adults in the UK. JAMA Netw Open, 2023. 6(3): p. e234714.

17. Satija, A., et al., Plant-Based Dietary Patterns and Incidence of Type 2 Diabetes in US Men and Women: Results from Three Prospective Cohort Studies. PLoS Med, 2016. 13(6): p. e1002039.

18. Willett, W.C., G.R. Howe, and L.H. Kushi, Adjustment for total energy intake in epidemiologic studies. Am J Clin Nutr, 1997. 65(4 Suppl): p. 1220S–1228S; discussion 1229S-1231S.

19. Fried, L.P., et al., Frailty in Older Adults: Evidence for a Phenotype. The Journals of Gerontology: Series A, 2001. 56(3): p. M146–M157.

20. Hanlon, P., et al., Frailty and pre-frailty in middle-aged and older adults and its association with multimorbidity and mortality: a prospective analysis of 4930737 UK Biobank participants. The Lancet Public Health, 2018. 3(7): p. e323–e332.

21. Pradhananga, S., et al., Ethnic differences in the prevalence of frailty in the United Kingdom assessed using the electronic Frailty Index. AGING MEDICINE, 2019. 2(3): p. 168–173.

22. Demirdağ, F., et al., Nutritional status as a mediator between the age-related muscle loss and frailty in community-dwelling older adults. Archives of Gerontology and Geriatrics, 2022. 98: p. 104569.

23. Hoogendijk, E.O., et al., Explaining the association between educational level and frailty in older adults: results from a 13-year longitudinal study in the Netherlands. Annals of Epidemiology, 2014. 24(7): p. 538-544.e2.

24. Kehler, D.S. and O. Theou, The impact of physical activity and sedentary behaviors on frailty levels. Mechanisms of Ageing and Development, 2019. 180: p. 29–41.

25. Kojima, G., et al., A systematic review and meta-analysis of prospective associations between alcohol consumption and incident frailty. Age and Ageing, 2017. 47(1): p. 26–34.

26. Webster, J., et al., Nutritional strategies to optimise musculoskeletal health for fall and fracture prevention: Looking beyond calcium, vitamin D and protein. Bone Rep, 2023. 19: p. 101684.

27. Frampton, J., et al., Higher dietary fibre intake is associated with increased skeletal muscle mass and strength in adults aged 40 years and older. Journal of Cachexia, Sarcopenia and Muscle, 2021. 12(6): p. 2134–2144.

28. Struijk, E.A., et al., Protein intake and risk of frailty among older women in the Nurses’ Health Study. Journal of Cachexia, Sarcopenia and Muscle, 2022. 13(3): p. 1752–1761.

29. Foster, H.M.E., et al., The effect of socioeconomic deprivation on the association between an extended measurement of unhealthy lifestyle factors and health outcomes: a prospective analysis of the UK Biobank cohort. The Lancet Public Health, 2018. 3(12): p. e576–e585.

30. Feraco, A., et al., Assessing gender differences in food preferences and physical activity: a population-based survey. Front Nutr, 2024. 11: p. 1348456.

31. Lewer, D., et al., Unravelling the alcohol harm paradox: a population-based study of social gradients across very heavy drinking thresholds. BMC Public Health, 2016. 16(1): p. 599.

32. Lana, A., F. Rodriguez-Artalejo, and E. Lopez-Garcia, Dairy Consumption and Risk of Frailty in Older Adults: A Prospective Cohort Study. Journal of the American Geriatrics Society, 2015. 63(9): p. 1852–1860.

33. Lorenzo-López, L., et al., Nutritional determinants of frailty in older adults: A systematic review. BMC Geriatrics, 2017. 17(1): p. 108.

34. Ma, Y., et al., Number of 24-Hour Diet Recalls Needed to Estimate Energy Intake. Annals of Epidemiology, 2009. 19(8): p. 553–559.

35. Struijk, E.A., et al., Sweetened beverages and risk of frailty among older women in the Nurses’ Health Study: A cohort study. PLOS Medicine, 2020. 17(12): p. e1003453.

36. Otten, D., et al., Similarities and Differences of Mental Health in Women and Men: A Systematic Review of Findings in Three Large German Cohorts. Frontiers in Public Health, 2021. 9.

37. Crimmins, E.M., J.K. Kim, and A. Solé-Auró, Gender differences in health: results from SHARE, ELSA and HRS. European Journal of Public Health, 2010. 21(1): p. 81–91.

38. Canon, M.E. and E.M. Crimmins, Sex differences in the association between muscle quality, inflammatory markers, and cognitive decline. The Journal of nutrition, health and aging, 2011. 15(8): p. 695–698.

39. Shivappa, N., et al., The Relationship Between the Dietary Inflammatory Index and Incident Frailty: A Longitudinal Cohort Study. Journal of the American Medical Directors Association, 2018. 19(1): p. 77–82.

40. Fry, A., et al., Comparison of Sociodemographic and Health-Related Characteristics of UK Biobank Participants With Those of the General Population. Am J Epidemiol, 2017. 186(9): p. 1026–1034.

